# The Impact of Frequent SARS-CoV-2 Testing on Weekly Case Rates Among Long-Term Care Facilities in Florida

**DOI:** 10.1101/2022.01.18.22268929

**Authors:** Lao-Tzu Allan-Blitz, Belal Aboabdo, Isaac Turner, Jeffrey D. Klausner

**Affiliations:** Division of Global Health Equity: Department of Medicine, Brigham and Women’s Hospital, Boston, MA; Curative Inc. San Dimas, CA; Departments of Preventive Medicine and Medicine, Keck School of Medicine, University of Southern California, Los Angeles, CA

## Abstract

SARS-CoV-2 testing frequency may be as important as test performance for disease control. We analyzed 1,292,165 SARS-CoV-2 test results among 361 long-term care facilities across Florida after implementing twice monthly testing (June 2020-April 2021). Our findings demonstrate that an increase in testing frequency reduced weekly case rates.

## Introduction

The SARS-CoV-2 pandemic has affected various populations differently. Most notably is the high rate of mortality among residents of long-term care facilities and nursing homes. Of deaths due to SARS-CoV-2 in the United States, 31% have been linked to long-term care facilities, despite constituting only 4% of cases overall (1). The increased mortality in such populations is likely driven by several factors; predominantly, residents of long-term care facilities are more likely to be older and have chronic medical conditions. Both of those factors may predispose individuals to mortality from SARS-CoV-2 infection (2). Additionally, residents of long-term care facilities live in close proximity, thus facilitating transmission (3). Residents of long-term care facilities, therefore, are particularly vulnerable to SARS-CoV-2 infection and consequent mortality.

One strategy for controlling infection in congregate living facilities is frequent testing, with resultant isolation of those with positive test results, thus limiting the transmission potential of the virus. A report from a nursing home in Pennsylvania documented the potential benefits of that approach. Rapid containment of an outbreak of SARS-CoV-2 infection in that 135-bed facility was achieved through rapid and frequent serial testing every 3-5 days of all residents and staff (4). Routine testing is particularly important to identify pre-symptomatic and asymptomatic cases, which are thought to account for up to 40% of new infections (5) and contribute significantly to transmission. A modeling study concluded that increased frequency of testing among nursing home residents and staff may be sufficient to reduce outbreaks (6).

In Florida, the Department of Emergency Management, in conjunction with Governor DeSantis, mandated twice monthly SARS-CoV-2 testing for all employees of skilled nursing facilities, elder care facilities, and assisted living facilities (7) beginning June 7^th^ 2020, with many facilities implementing testing in June. For the duration of the program, only staff members were permitted entry to the facilities, and all visitors were barred from entry. Thus, we aimed to evaluate the impact of frequent testing on weekly SARS-CoV-2 case rates in a real-world setting.

## Methods

The initiative implemented by the Florida Department of Emergency Management included twice monthly testing for all employees of long-term care facilities in Florida. Curative Inc. sent mucosal oral fluid swab test collection kits to 361 facilities along with video instructions with details of sample collection for employees of the facilities to review.

We analyzed de-identified data for the 41-week period between June 2020 to April 2021 from 361 nursing homes. Trained clinical nursing home staff collected oral mucosal swab specimens according to the assay’s instructions for use (8). Specimens were transported in RNA preservative media (DNA/RNA Shield™ solution, Zymo Research Corp., Irvine, CA). The specimens were subsequently tested by a modified Centers for Disease Control and Prevention protocol using polymerase chain reaction, as has been previously described (9). Results were sent directly to the employee as well as to the facility and Florida Department of Public Health for contact tracing and program monitoring. Data collected included sample collection date and result, location of testing facility, report of any symptoms from the individual being tested. Data were not collected on the interventions taken by the individual facility upon receipt of a positive test result.

We then aggregated our dataset with the Nursing Home Provider Information data made publicly available by the Centers for Medicare & Medicaid Services (10), which included number of beds within a facility, number of facility staff, and average aid hours per resident. Our dataset was further enriched by joining time-series data, tracked by Johns Hopkins University, of daily SARS-CoV-2 cases indexed by day and county with data on rates of hospitalization and mortality (11). We report the average weekly cases, tests per occupied bed and new cases per 100,000 individuals as well as the standard deviations (SDs).

We used a generalized linear mixed regression model with weekly cases modeled as a negative binomial random count variable to assess how the independent variables impacted test positivity. We regressed weekly positive cases on the date twice-per-month testing began, cases from the preceding week, new cases per 100,000 people in the county, total tests per occupied bed (a surrogate for compliance with twice-per-month testing), number of certified beds in the facility, total nurse staffing hours per resident per day (as a surrogate for quality of care), and date of infection before or after January 2021 to control for any impact of vaccination. All variables were log transformed except cases in the week preceding. We applied a random effect for each nursing home. All analyses were performed using R statistical software (R Core Team, 2020).

## Results

During the study period, we analyzed 1,292,165 SARS-CoV-2 RNA test results collected from residents and employees from 361 facilities in Florida. The facilities were located across 247 zip codes. The average age of the study population was 49 years (SD +/-31). The average number of new cases among all long-term care facilities in Florida was 187.9 per week (SD +/-148.7), with an estimated 0.7 tests per week per occupied bed (SD +/-16.2). The average test turnaround time from laboratory receipt was 17.1 hours (SD +/-10.4). The average tests completed per week was 31,454 (SD +/-10,926).

We found a notable reduction in predicted SARS-CoV-2 case counts with increased testing in the preceding weeks, even when controlling for county-level prevalence, facility size, and daily aid hours per resident (see Table). We found that a 10% increase in testing frequency would result in a 1% reduction in weekly long-term care facility case rates among residents. Extrapolated, that reduction would result in 126 fewer cases per week among residents of long-term care facilities across the United States, or 6,552 fewer cases per year.

**Table.** Estimated Change in Weekly SARS-CoV-2 Cases due to Increased Testing Frequency Among Florida Long-Term Care Facilities June 2020 – April 2021

|  | **Change in Weekly**<br>**Cases** | **p-value** |
| --- | --- | --- |
| *Onset of Two times per month Testing\** |  |  |
| One week preceding | 0.10 (0.03 – 0.16) | 0.003 |
| Two weeks preceding | -0.02 (-0.09 – 0.04) | 0.53 |
| Three weeks preceding | -0.08 (-0.14 – -0.02) | 0.01 |
| Four weeks preceding | -0.03 (-0.09 – -0.03) | 0.34 |
| *Nursing home size and quality* |  |  |
| Certified beds per facility | 0.006 (0.004 – 0.008) | <0.001 |
| Aid hours per resident per facility | -0.07 (-0.17 – 0.03) | 0.16 |
| *Virus conditions* |  |  |
| Cases among long-term care facilities in preceding week | 0.09 (0.09 – 0.10) | <0.001 |
| New county-level cases per 100,000 individuals | 0.002 (0.001 – 0.002) | <0.001 |
| *Qualitative Variables* |  |  |
| Tested after January 2021<sup>†</sup> | -0.70 (-0.82 – -0.58) | <0.001 |

## Discussion

We analyzed SARS-CoV-2 testing data among 361 nursing homes in Florida and linked those data with national level registry data to evaluate the impact of routine twice-monthly testing on the weekly case rate. We adjusted our findings for county-level incidence, size of the nursing home, and a surrogate marker for quality of care. Overall, we noted a significant decrease in the incidence of SARS-CoV-2 infection among nursing homes adherent to twice-monthly testing of employees.

In our study, the impact on case rate was observed only after a one-week delay from the onset of increased testing frequency. That finding likely reflects multiple factors; predominantly that the incubation period of the virus is estimated to be on average six days (12) and thus the ability to detect the virus upon entry into a host will be similarly delayed. Additionally, from the moment of case identification, isolation of infected individuals and contact tracing require further time to initiate, thus a notable impact on reducing viral transmission will not be seen immediately. However, our findings are in agreement with prior modeling work concluding frequent testing can be an effective control strategy (6, 13), and support the use of routine frequent testing among residents and staff of long-term care facilities.

Importantly, the impact of repeated testing was seen even when controlling for the community prevalence of SARS-CoV-2 infection, thereby accounting for the potential confounding of reducing county-level prevalence as an explanation for our results. The specific frequency of testing, however, will likely be determined at least in part by community level prevalence (14), with our results from twice monthly testing perhaps not generalizable across counties with varying SARS-CoV-2 prevalence.

We also found that increased aid time with residents was associate with a small reduction the weekly SARS-CoV-2 case rate. That finding is consistent with prior work (15). It is possible that such increased time resulted in earlier identification of more mild symptoms, and thus reduced transmission opportunities. However, that potential protective effect may only be evident in populations where staff are tested regularly, thereby mitigating the risk of transmission from staff to residents.

## Limitations

Our study has several limitations. Overall, because we do not have specific details on the interventions implemented as a result of increased testing frequency, we cannot directly attribute the above findings solely to the increased frequency of testing. Additionally, our study was from a single state, thus generalizability may be limited. The strengths of our study, however, are the large sample size, increasing the precision of our findings, and the numerous sites within Florida that were included.

## Conclusion

Our results indicate that twice-monthly testing for SARS-CoV-2 among employees of long-term care facilities was associated with reduced case rates even when controlling for county prevalence, size of the facility, and quality of care delivered. Our findings support the practice of routine, frequent testing among long-term care facilities, with the specific interval of testing guided by county prevalence.

## Data Availability

Data are available upon request

## Acknowledgements and Funding

The authors would like to acknowledge Mitchel Roznik for his contributions to the project as well as the State of Florida. There was no funding for this project. This research was supported in part by a gift to the Keck School of Medicine of the University of Southern California by the W.M. Keck Foundation

## References

1. New York Times. Nearly One-Third of U.S. Coronavirus Deaths Are Linked to Nursing Homes. Updated June 1, 2021. https://www.nytimes.com/interactive/2020/us/coronavirus-nursing-homes.htm Accessed June 10, 2021.

2. Liu K, Chen Y, Lin R, Han K. Clinical features of COVID-19 in elderly patients: A comparison with young and middle-aged patients. J Infect. 2020;80(6):e14–e8.

3. Cerami C, Rapp T, Lin FC, et al. High household transmission of SARS-CoV-2 in the United States: living density, viral load, and disproportionate impact on communities of color. medRxiv. 2021.

4. Escobar DJ, Lanzi M, Saberi P, et al. Mitigation of a COVID-19 Outbreak in a Nursing Home Through Serial Testing of Residents and Staff. Clin Infect Dis. 2020.

5. Louie JK, Scott HM, DuBois A, et al. Lessons From Mass-Testing for Coronavirus Disease 2019 in Long-Term Care Facilities for the Elderly in San Francisco. Clin Infect Dis. 2021;72(11):2018–20.

6. Holmdahl I, Kahn R, Hay JA, Buckee CO, Mina MJ. Estimation of Transmission of COVID-19 in Simulated Nursing Homes With Frequent Testing and Immunity-Based Staffing. JAMA Netw Open. 2021;4(5):e2110071.

7. Agency for Health Care Administration: Florida. Rule 59AER20-5 Mandatory Testing for Nursing Home Staff. Available at: https://ahca.myflorida.com/docs/59AER20-5_Mandatory_Testing_for_Nursing_Home_Staff.pdf Accessed June 22, 2021.

8. DNA/RNA ShieldTM Collection Tube w/Swab. Package Insert. Available at: https://files.zymoresearch.com/pdf/r1106i.pdf Accessed June 14, 2021.

9. Kojima N, Turner F, Slepnev V, et al. Self-Collected Oral Fluid and Nasal Swab Specimens Demonstrate Comparable Sensitivity to Clinician-Collected Nasopharyngeal Swab Specimens for the Detection of SARS-CoV-2. Clin Infect Dis. 2020.

10. Centers for Medicare Services Data. COVID-19 Nursing Home Data. Available at: https://data.cms.gov/stories/s/COVID-19-Nursing-Home-Data/bkwz-xpvg/ xLast Updated May 16, 2021. Acccessed June 1, 2021.

11. Dong E, Du H, Gardner L. An interactive web-based dashboard to track COVID-19 in real time. Lancet Infect Dis. 2020;20(5):533–4.

12. Wassie GT, Azene AG, Bantie GM, Dessie G, Aragaw AM. Incubation Period of Severe Acute Respiratory Syndrome Novel Coronavirus 2 that Causes Coronavirus Disease 2019: A Systematic Review and Meta-Analysis. Curr Ther Res Clin Exp. 2020;93:100607.

13. See I, Paul P, Slayton RB, et al. Modeling effectiveness of testing strategies to prevent COVID-19 in nursing homes - United States, 2020. Clin Infect Dis. 2021.

14. Chin ET, Huynh BQ, Chapman LAC, Murrill M, Basu S, Lo NC. Frequency of Routine Testing for Coronavirus Disease 2019 (COVID-19) in High-risk Healthcare Environments to Reduce Outbreaks. Clin Infect Dis. 2020.

15. Figueroa JF, Wadhera RK, Papanicolas I, et al. Association of Nursing Home Ratings on Health Inspections, Quality of Care, and Nurse Staffing With COVID-19 Cases. JAMA. 2020.

